# Initial emergency department vital signs may predict PICU admission in pediatric patients presenting with asthma exacerbation

**DOI:** 10.1101/2021.09.22.21263918

**Authors:** Michael S Freedman, Erick Forno

**Author notes:** Corresponding Author: Michael S Freedman, MD MPhil.

## Abstract

**Objective:** Severe asthma exacerbations account for a large share of asthma morbidity, mortality, and costs. Here, we aim to identify early predictive factors for pediatric intensive care unit (PICU) admission that could help improve outcomes.

**Methods:** We performed a retrospective observational study of 6,014 emergency department (ED) encounters at a large children’s hospital, including 95 (1.6%) resulting in PICU admission between 10/1/2015 and 8/31/2017 with ICD9/ICD10 codes for “asthma,” “bronchospasm,” or “wheezing”. Vital signs and demographic information were obtained from EHR data and analyzed for each encounter. Predictive factors were identified using adjusted regression models, and our primary outcome was PICU admission.

**Results:** Higher mean heartrates (HR) and respiratory rates (RR) and lower SpO2 within the first hour of ED presentation were independently associated with PICU admission. Odds of PICU admission increased 63% for each 10-beats/minute higher HR, 97% for each 10-breaths/minute higher RR, and 34% for each 5% lower SpO2. A binary predictive index using 1-hour vitals yielded OR 11.7 (95%CI 7.4-18.3) for PICU admission, area under the receiver operator characteristic curve (AUROC) 0.82 and overall accuracy of 81.5%. Results were essentially unchanged (AUROC 0.84) after adjusting for asthma severity and initial ED management. In combination with a secondary standardized clinical asthma distress score, positive predictive value increased by seven-fold (5.9% to 41%).

**Conclusions:** A predictive index using HR, RR and SpO2 within the first hour of ED presentation accurately predicted PICU admission in this cohort. Automated vital signs trend analysis may help identify vulnerable patients quickly upon presentation.

## INTRODUCTION

Asthma affects over 330 million people worldwide (1). In the U.S. alone, it leads to 1.8 million emergency department (ED) visits and 189 000 hospital admissions every year (2), with annual costs estimated in excess of $81B (3). Every year, over 3500 people die from asthma in the U.S. (2) While outcomes have generally improved, we lack early predictors of need for care escalation in pediatric patients with acute asthma exacerbations presenting to the ED. Beyond general clinical guidelines (4,5), treatment protocols vary across institutions, resulting in equally variable clinical outcomes.

Prior studies to predict asthma outcomes have analyzed outpatient data on air quality (6) to Twitter trends (7) to the volume of ED visits for asthma. Medication regimens and vital signs have also been used to predict hospital admission rates of children with asthma exacerbations (8). To date, however, there is limited data on specific risk factors for PICU admission, with a handful of studies focusing on demographic (9) or environmental factors (10) and signs of impending clinical deterioration such as cyanosis (8,11,12). Scoring systems like the pediatric asthma score (PAS), the modified pulmonary index score (MPIS), or the asthma distress score (ADS) combine clinical signs of exacerbation severity and have been validated to predict hospitalizations (13,14), PICU admissions (15), and length of stay (LOS) (16); however, they require subjective assessments that can vary widely between examiners (e.g. accessory muscle use, location and quality of breath sounds, etc.).

Objective approaches derived from clinical and objective measurements may correlate with some of these scoring systems (17), but to date they have not been validated to predict PICU admissions. Timely identification of patients at high risk of PICU admission and increased LOS is critical to expedite care, escalate treatment, reduce morbidity, and reduce burden on both the hospital system and the family.

In this study, we aim to identify risk factors for PICU admission and increased LOS. We focus on early and objective measures that could be incorporated into the development of automated clinical algorithms using the electronic health record (EHR), with the goal of improving efficiency and reducing provider variability.

## METHODS

### Study population and data extraction

EHR data were extracted for all ED encounters at our institution for the period between 10/1/2015 and 8/31/2017, using ICD9 and ICD10 codes for “asthma,” “bronchospasm,” or “wheezing”. Variables included demographic information (age, sex, reported race/ethnicity); height; weight; all vital signs (heart rate [HR], respiratory rate [RR], and O2 saturations [SpO2]) as well as Asthma Distress Scores [ADS] throughout the ED visit; and the timeframe for presentation, admission, transfer, and discharge orders. Management data included albuterol doses; use of steroids, magnesium sulfate, and epinephrine; and controller medications prescribed during the admission (including inhaled corticosteroids [ICS] and long-acting beta agonists [LABA]). The study was approved by the UPMC Quality Improvement (QI) Review Committee (project ID #1581), and consent was waived given the retrospective nature of the EHR review.

Vital signs were summarized by calculating the distribution of measurements for each patient the first 60 minutes in the ED: for each patient, we calculated mean, standard deviation, median, and slope for HR, RR, and SpO2. These summary variables were used as potential predictors in our analysis.

### Outcomes and statistical analysis

Our primary outcome was PICU admission. Secondary outcomes were ED, PICU, and total LOS, calculated based on the interval between ED presentation and admission, transfer, or discharge orders. Potential predictors for PICU admission included average HR, RR, and SpO2 within the first hour of presentation to ED and medication interventions. First, we analyzed the association between each potential predictor separately and the outcome of interest, using regression models (logistic for PICU admission or linear for LOS) adjusted for age, sex, and reported race/ethnicity. Predictors that were significantly associated with the outcome were then grouped in one model, and those that remained significant (along with age and sex) were used to construct a 1-hour vital signs predictive index (hereinafter “VSPI”), where PICU_pred_ is the likelihood of PICU admission, and β describes the effect size of each variable:

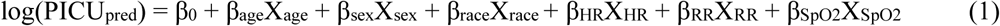

The VSPI was compared to actual PICU admission using receiver-operating characteristics (ROC), and then analyzed to determine the optimal cut-off point for a binary index. The performance of the binary index was determined by assessing the area under the ROC curve (AUROC) and its accuracy, sensitivity, specificity, negative and positive predictive values (NPV and PPV, respectively), and overall accuracy (true positive plus true negatives / whole sample). A similar predictive index using ADS and demographics was constructed for comparison.

We proceeded in a similar fashion using linear regression models to examine ED, PICU, and overall LOS, in the whole sample and stratified by disposition (i.e., hospitalized, PICU, and discharged from the ED):

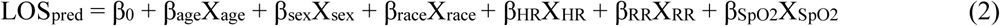

Where LOS_pred_ describes predicted length of stay in hours, and β describes the effect size for each variable. All analyses were performed in RStudio version 1.2.5042, and a two-sided P < 0.05 was considered as statistically significant. Analyses were performed on encounters for which complete data were available, omitting encounters with incomplete data without imputation. Study was developed in accordance with TRIPOD and RECORD checklists, and model performance measures were generated in accordance with current guidelines (18).

## RESULTS

Among 6,251 inpatient encounters for asthma exacerbation, 6208 (99.3%) presented via the ED and 6014 (96.2%) had a complete set of initial vital signs available; 95 (1.6%) of these resulted in PICU admission (Figure 1). Baseline encounter characteristics are shown in Table 1. Initial vital signs for patients who were admitted to the PICU showed higher HR, higher RR, and lower SpO_2_ compared to those who were not admitted to the PICU (Table 1). Patients who required PICU admission were more likely to be white, to have received magnesium sulfate, epinephrine, or high-dose continuous albuterol in the first hour of ED presentation, and to have prescriptions for ICS or LABA. There were no significant differences in age or sex between patients who were admitted to the PICU and those who were not.

**Table 1.**
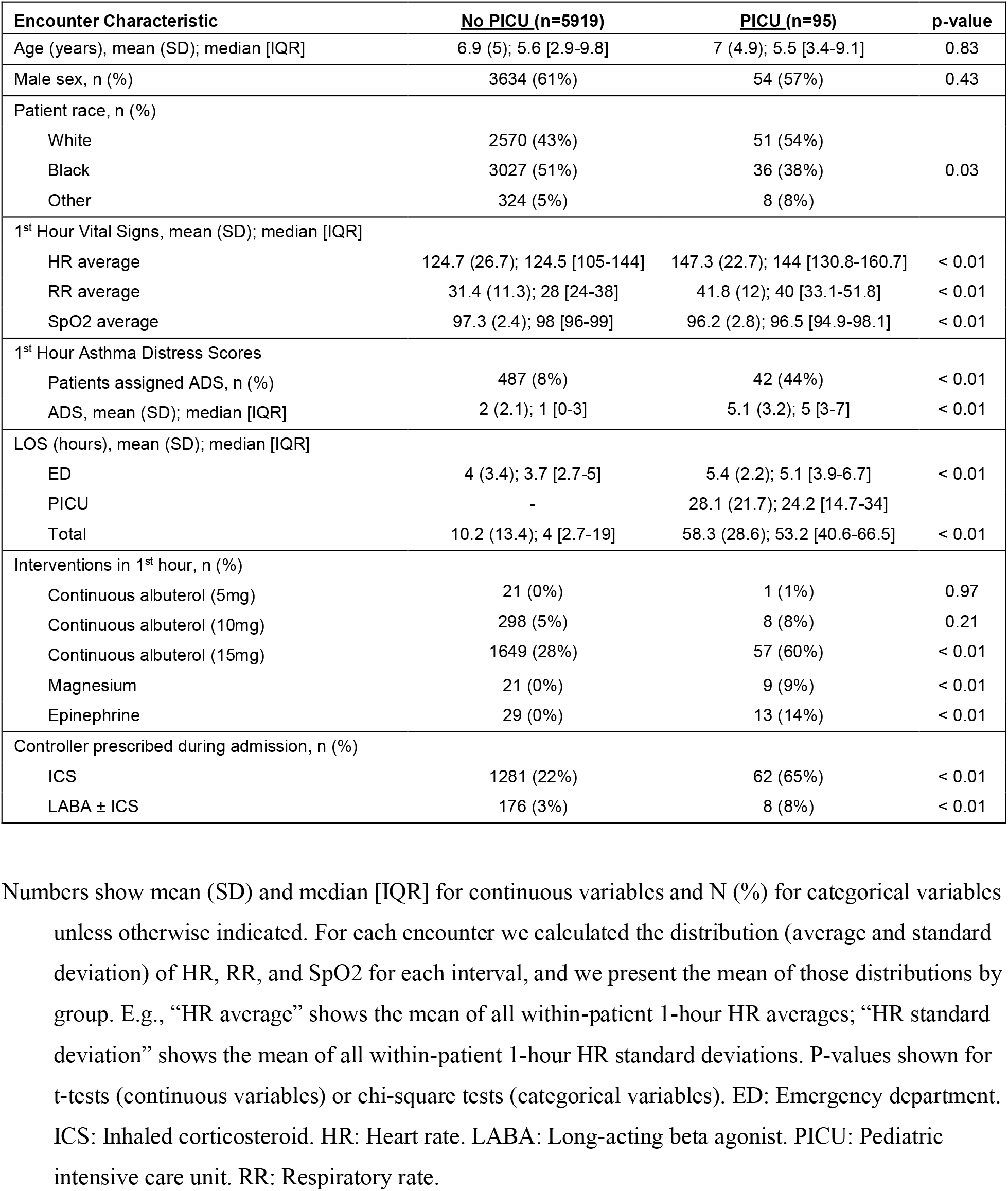
Baseline encounter characteristics

**Figure 1.**
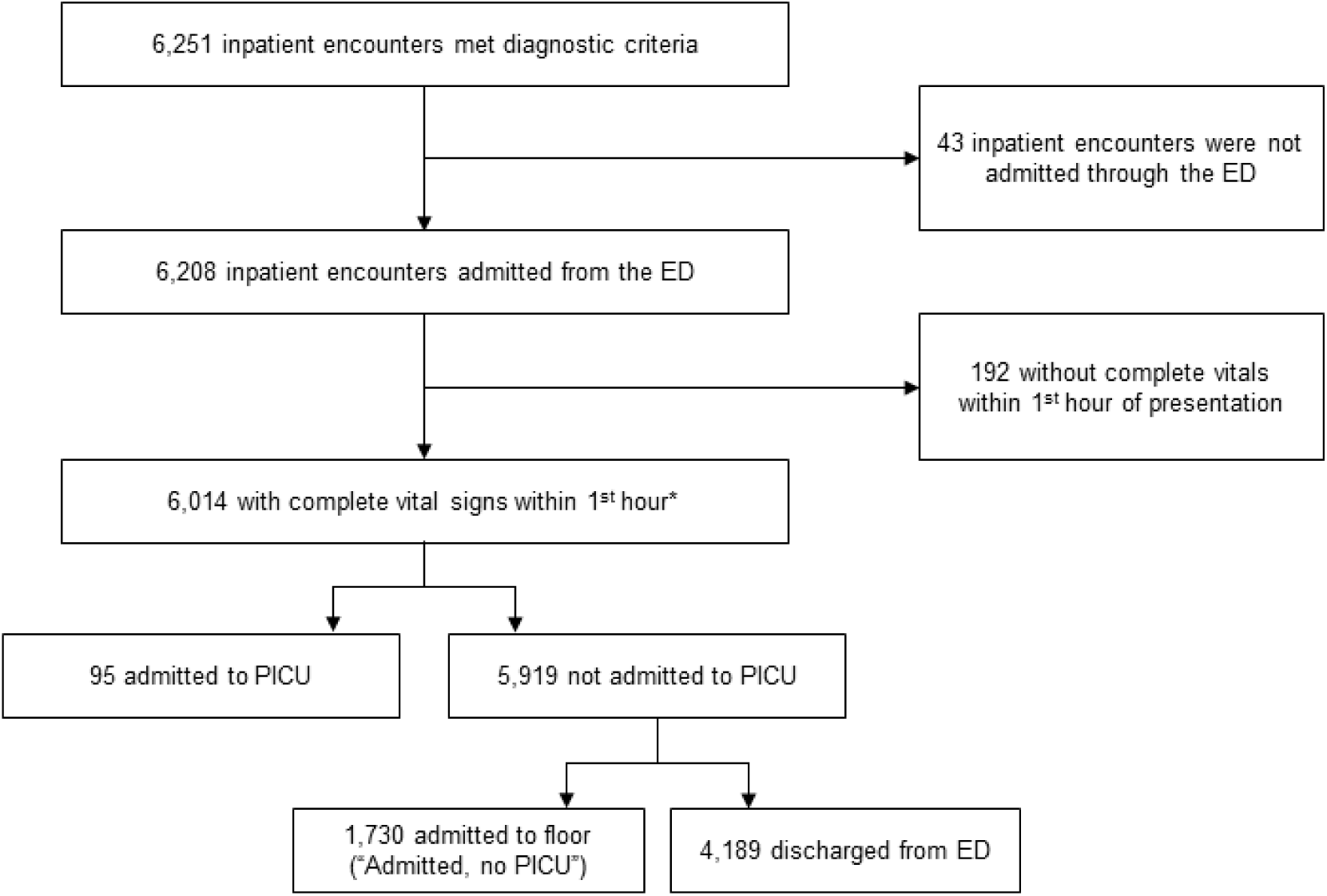
Flowchart of study selection. Encounters that met inclusion criteria and admitted to PICU vs not admitted to PICU. *2 encounters excluded due to erroneous data (mis-specified discharge date/time).

### PICU admission

Vital signs within the first hour in the ED were associated with PICU admission after adjusting for age, sex, and race. Higher average HR (Table 2, <u>Model 1</u>: OR 1.05, 95%CI 1.03-1.06) and higher average RR (Table 2, <u>Model 2</u>: OR 1.07, 95%CI 1.06-1.09) were each associated with increased odds of PICU admission: a 10-bpm higher HR was associated with 63% increased odds of PICU admission, while 10 breaths-per-minute higher RR was associated with 97% increased odds of PICU admission. Similarly, average SpO2 within the first hour of ED presentation was inversely associated with the odds of PICU admission (Table 2, <u>Model 3</u>: OR 0.92, 95%CI 0.88-0.96): each 5% drop in SpO2 was associated with 47% increased odds of PICU admission. HR, RR, and SpO_2_ remained independently associated when incorporated into the same model (Table 2, <u>Model 4</u>).

**Table 2.** Adjusted regression models for PICU admission

| Encounter<br>Characteristic | Model 1<br>OR (95%CI), p-value | Model 2<br>OR (95%CI), p-value | Model 3<br>OR (95%CI), p-value | Model 4<br>OR (95%CI), p-value | Model 5<br>OR (95%CI), p-value |
| --- | --- | --- | --- | --- | --- |
| Age (years) | **1.14 (1.10 - 1.19), < 0.01** | **1.10 (1.06 - 1.13), < 0.01** | 1.02 (0.98 - 1.06), 0.40 | **1.16 (1.11 - 1.21), < 0.01** | 1.03 (1.00 - 1.06), 0.09 |
| Male sex | 1.00 (0.66 - 1.54), 0.98 | 0.85 (0.56 - 1.3), 0.47 | 0.81 (0.54 - 1.23), 0.33 | 1.00 (0.65 - 1.54), 0.99 | 0.93 (0.61 - 1.42), 0.75 |
| Patient race |  |  |  |  |  |
| Black (vs White) | 0.71 (0.46 - 1.11), 0.14 | **0.58 (0.37 - 0.91), 0.02** | **0.62 (0.4 - 0.96), 0.03** |  |  |
| Other (vs White) | 1.10 (0.51 - 2.38), 0.81 | 0.98 (0.45 - 2.14), 0.96 | 1.25 (0.59 - 2.67), 0.56 |  |  |
| 1-Hour Vitals |  |  |  |  |  |
| HR average | **1.05 (1.03 - 1.06), < 0.01** |  |  | **1.03 (1.02 - 1.04), < 0.01** |  |
| RR average |  | **1.07 (1.06 - 1.09), < 0.01** |  | **1.05 (1.03 - 1.07), < 0.01** |  |
| SpO2 average |  |  | **0.92 (0.88 - 0.96), < 0.01** | **0.94 (0.89 - 0.99), 0.02** |  |
| **Predictive Index** |  |  |  |  | **2.72 (2.26 - 3.27), < 0.01** |

Adjusted logistic regression models examining the effects of demographics and initial vital signs independently (Models 1-3) and incorporated into the same model (Model 4) to develop a continuous predictive index (Model 5).

Given our interest on early prediction, we created a composite index using the effect sizes from that model (Table 2, <u>Model 5</u>), which in our data ranged from −2.35 to +6.07 and had different distributions for PICU and no PICU (Figure 2). Each 1-point increase in the predictive index was associated with 2.7-fold higher odds of PICU admission (OR 2.7, 95%CI 2.3-3.3), and the index had an AUROC of 82%. Using Youden’s index, we selected a cut-off value of 1.06; with this cut-off, the binary index yielded an odds ratio of 11.7 (95%CI 7.4-18.3) for PICU admission, with sensitivity 72%, specificity 82%, NPV 99%, PPV 5.9%, and overall accuracy of 81.5% (Figure 2). Results remained essentially unchanged after additionally adjusting for initial ED management, including administration of magnesium sulfate, epinephrine, or continuous albuterol (<u>Supplemental Figure 1</u>; AUROC=0.84, sensitivity 80%, specificity 77%, NPV 99.6%, PPV 5.3%, overall accuracy 77.2%), albuterol dose (AUROC=0.83, sensitivity 80%, specificity 76%, NPV 99.6%, PPV 5.0%, overall accuracy 75.7%), or whether the patient was prescribed an ICS or LABA (AUROC=0.87, sensitivity 84%, specificity 75%, NPV 99.7%, PPV 5.1%, overall accuracy 74.8%).

**Figure 2.**
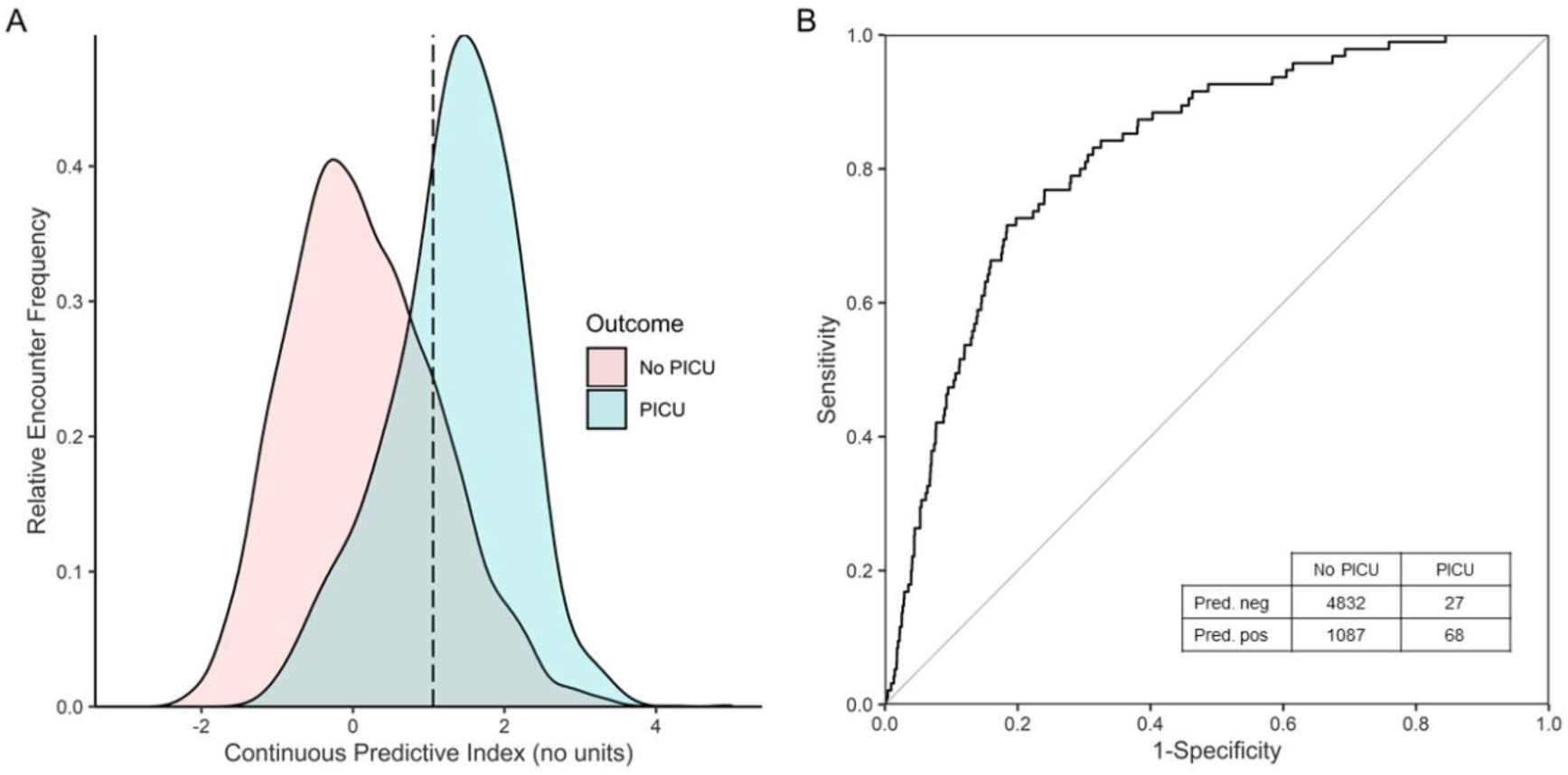
One-hour vital sign predictive index and PICU admission. (A) Continuous 1-hour predictive index vs PICU admission, showing the optimal binary threshold = 1.06 (dotted line). (B) Receiver-Operator Curve (ROC) and classification table of 1-hour predictive index vs PICU admission. AUROC=0.83, sensitivity 72%, specificity 82%, negative predictive value 99.4%, and positive predictive value 5.9%. Overall accuracy 81.5% (true positives plus true negatives as percent of total).

We then compared the VSPI to the asthma distress score (ADS), a clinical score of asthma severity used in our institution, similar to other validated asthma severity scores (19–21); in addition to RR and SpO_2_, the ADS requires physical examination to determine auscultatory findings, the degree of dyspnea, and the use of accessory muscles. Data to calculate the ADS within the first hour of presentation were available for 529 (8.8%) of ED encounters, including 42 resulting in PICU admission. Among those 529 encounters, the ADS yielded similar performance overall but had lower sensitivity than the VSPI: AUROC=0.81, sensitivity 21.4%, specificity 92%, NPV 93%, PPV 18%, overall accuracy 86%. In that same subset of 529 encounters, our 1-hour VSPI had AUROC=0.82, sensitivity 81%, specificity 66%, NPV 98% and PPV 17%. A similarly structured predictive index using ADS (ADSPI) was then applied as a second-tier to the 210 encounters that initially screened positive by VSPI. Of these, 152 were ADSPI-negative with 12 PICU admissions, and 58 were ADSPI-positive with 24 PICU admissions (AUROC=0.79, sensitivity 67%, specificity 80%, NPV 92% PPV 41%). Using both as a two-step predictive index (TSPI) yielded sensitivity 57%, specificity 93%, NPV 96% and PPV 41% (Table 3).

### Length of stay

Among patients admitted to the PICU, the average ED, PICU, and total LOS were 5.4 hours (95%CI 1.9–9.8), 28.1 hours (95%CI 6.5–98.0), and 58.3 hours (95%CI 26.2–143.8), respectively (Table 1). Among those who were not admitted to the PICU, average ED and total LOS were 4.0 hours (95%CI 1.3–8.4) and 10.4 hours (95%CI 1.3–40.1), respectively. And in the subset of encounters in which patients were discharged directly from the ED, average LOS was 3.4 hours (95%CI 1.2–6.6).

Among patients admitted to the PICU (n=95), none of the included characteristics were associated with either PICU LOS or total LOS (<u>Supplemental Table 1</u>). Furthermore, adjusted models for children admitted to the PICU showed significant but only weak associations with PICU LOS (R^2^=0.02; <u>Supplemental Figure 2A</u>) and total LOS (R^2^=0.07; <u>Supplemental Figure 2B</u>). Among patients who were admitted to the hospital but not the PICU (n=1,730), we found significant but weak associations between older age, higher average HR, and lower average SpO2, and longer total LOS. Multivariable models explained only a small proportion of the variability in total hospital LOS (R^2^=0.05; <u>Supplemental Table 2</u>). The addition of further variables to the model, including additional therapies and longer time windows, resulted in higher but still very modest R^2^ values (<u>Supplemental Table 2</u>).

## DISCUSSION

In our analysis, we report an index composed of heart rate, respiratory rate, and SpO_2_ that was highly predictive of PICU admission among children presenting with asthma to the ED. While computerized scoring systems have been previously developed to standardize asthma severity, to our knowledge this analysis demonstrates the first predictive model that can be fully automated and anticipate escalation of care to the ICU.

The difficulty in adequately triaging severe asthma in the ED is multifaceted. A changing healthcare landscape places increased utilization and stress on emergency departments, including pediatric emergency departments, producing an influx of patients on an already strained system of limited providers and resources. This is particularly important in the setting of long-term asthma incidence that has been steadily increasing, with worsening environmental factors exacerbated by climate change (22–25), which can in turn lead to more frequent and more severe asthma exacerbations. Moreover, the frequency of pediatric asthma exacerbations severe enough to require escalation to PICU-level care may be low (1.6% in our analysis), but the resulting morbidity in these patients – and the cost on the healthcare system – is high, highlighting the importance of a system to easily identify these few patients amongst the many.

Our predictive index has many strengths over more subjective, scoring-based triage systems. Using just demographic variables and the first 60 minutes of vital signs, the predictive index achieved an overall accuracy of ∼82%, and children with a positive index were 11 times more likely to require PICU than those with a negative index. Moreover, the accuracy of the index did not change after accounting for asthma severity and initial management in the ED. The predictive index had similar AUROC, overall accuracy, and positive predictive value compared to ADS-based triage; however, the proposed index has the benefit of relying on strictly objective data, it does not require active involvement by a provider who can perform a physical exam, and therefore it is computable for all patients with a full set of vital signs in the first hour (compared to the <10% of ED encounters with a full ADS in the first hour of presentation). Furthermore, as a patient’s clinical presentation develops and vital signs change, this information could be automatically monitored to refine the estimated risk of PICU admission.

The variables that were most strongly associated with PICU admission were higher average HR, followed by higher average RR, and finally lower average SpO2. Our approach integrates these variables into one index that can be easily automated. There were no strong associations between race and PICU admission, and while increased age was a risk factor in some of our models, it was not significantly associated with PICU admission when integrated into the combined model. It is conceivable that older children presenting with an asthma exacerbation may have a more severe phenotype with increased odds of PICU admission. Unsurprisingly, quick administration of magnesium or epinephrine was more common among ED encounters that ultimately resulted in PICU admission, but these interventions were excluded from the predictive index as they rely on clinical evaluation and judgment, and they would be invariably linked to disposition. Nonetheless, further adjusting the models by these variables did not substantively change the utility of the predictive index, further demonstrating the robustness of the approach.

As PICU admission for asthma is a low-frequency event, the VSPI has excellent negative predictive value, but its positive predictive value remains low even when measured across various levels of sensitivity (recall). By further evaluating VSPI+ patients using the asthma distress score predictive index (ADSPI), PPV improves by seven-fold to 41%, at minimal expense to NPV (96% from 99%). A two-step approach has multiple strengths. The VSPI can initially screen out the vast majority of asthmatics by vital signs and demographics alone due to its excellent NPV, with potential for full EHR automation. Those who screen positive can then be assessed clinically to identify patients at highest risk of PICU. Rapid stratification of low-risk patients (by VSPI) can likewise alleviate the burden of assigning ADS or similar scores to all children presenting with asthma.

In our cohort, patients admitted to the PICU experienced a longer length of stay (LOS) in the ED than those who were not admitted to the PICU. However, among patients admitted to the PICU, LOS (in the PICU or total) did not differ significantly by age, sex, race, or the initial vital signs. This is likely related to the heterogeneity in severe asthma phenotypes and their response to treatment; while vital signs and demographics may predict who may need higher level of care, they are less helpful in determining how they will respond or how long they will need to remain admitted. Among patients who avoided PICU admission, older age, male sex, white race, and similar vital sign changes suggestive of respiratory distress (tachycardia, tachypnea and hypoxia) were each associated with longer total LOS. While associations between race, sex and asthma severity are well documented (26,27), the association between longer total LOS, white race and male sex is not well understood. Altogether, however, our models were not helpful to predict PICU LOS and total LOS (Supplemental Figures 2A, 2B).

Our study had several limitations. This was a retrospective analysis based on data queried from the EHR using diagnostic codes. Our rate of PICU admission was low, and this limited our ability to detect small effect sizes or to perform specific subgroup analyses. Escalation of care to PICU for acute asthma exacerbation will likely differ across institutions, limiting the generalizability of our findings. At our institution, administration of respiratory support of high-flow nasal cannula or CPAP requires PICU; external validity may vary based on PICU admission criteria at different institutions. Finally, a nuanced understanding of vital sign changes is required if this index were to be implemented. Vital signs incorporated in the VSPI were not adjusted for temperature, as its measurement is source-dependent and not consistently accurate. HR and RR were elevated in both groups relative to baselines for age, requiring calibration of any potential future implementation to be normalized to the tachypnea and tachycardia associated with presenting to the ED in respiratory distress with confounding potential pain or anxiety.

At the same time, our study had several strengths worth noting. First, despite a relatively small number of PICU admissions, our analysis was based on an extensive dataset from a large referral center. Second, we had complete data for each encounter, and thus there was no significant loss of data in terms of the outcomes analyzed. A particular strength of our analysis is that the performance of the predictive index remained strong despite a noisy dataset, and despite adjustment for several covariates, including asthma severity and initial management in the ED. Finally, index performance was at least similar to the ADS, which requires substantially higher involvement to complete and is prone to inter-rater variability.

## Conclusions

We report an index that performs well in predicting PICU admissions among children presenting with asthma exacerbations to the ED. This index is easily obtained and could be automated as a prognostic aid for physicians taking care of these children. Future directions will include a prospective, independent validation of an automated version of this predictive index, as well as alternate machine-learning approaches to improve its performance.

## Data Availability

Data is available from authors upon request.

## Abbreviations

ADS: asthma distress score
ADSPI: asthma distress score predictive index
AUROC: area under receiver operator characteristic
ED: emergency department
EHR: electronic health record
HR: heart rate
LOS: length of stay
NPV: negative predictive value
PICU: pediatric intensive care unit
PPV: positive predictive value
RR: respiratory rate
SPO2: oxygen saturation
TSPI: two-step predictive index
VSPI: vital signs predictive index

