## Supplemental Materials for "Initial emergency department vital signs may predict PICU admission in pediatric patients presenting with asthma exacerbation"

### Supplemental Material

Supplemental Table 1. Adjusted regression models for PICU and total length of stay

| | | PICU (n=95) | | Admitted, No PICU (n=1,730) |
|---|---|---|---|---|
| | | **PICU LOS** | **Total LOS** | **Total LOS** |
| **Covariates** | **Regression Coefficient** | **Beta (SE), p-value** | **Beta (SE), p-value** | **Beta (SE), p-value** |
| Age (years) | $\beta_{age}$ | 0.14 (0.72), 0.84 | 1.30 (0.92), 0.16 | **0.83 (0.11), < 0.01** |
| Male sex (vs Female) | $\beta_{sex}$ | 1.87 (5.22), 0.72 | 0.87 (6.64), 0.9 | 1.00 (0.73), 0.17 |
| Black race (vs White) | $\beta_{race}$ | 2.11 (5.27), 0.69 | 8.69 (6.70), 0.2 | 0.38 (0.72), 0.59 |
| Vital Signs (1st Hour) | | | | |
| HR average | $\beta_{HR}$ | -0.01 (0.17), 0.96 | -0.02 (0.21), 0.93 | **0.05 (0.02), < 0.01** |
| RR average | $\beta_{RR}$ | 0.01 (0.26), 0.99 | 0.14 (0.33), 0.67 | 0.00 (0.04), 0.99 |
| SpO2 average | $\beta_{SpO2}$ | -0.96 (0.89), 0.29 | -1.74 (1.13), 0.13 | **-0.39 (0.14), < 0.01** |

HR: Heart rate. LOS: Length of stay. RR: Respiratory rate. SpO2: Oxygen saturation. SE: Standard error.

Supplemental Table 2. Adjusted regression models for total LOS among children admitted without PICU

| Encounter Characteristics | Model A | Model B | Model C | Model D |
|---|---|---|---|---|
| **Model $R^2$** | 0.05 | 0.05 | 0.06 | 0.06 |
| **Demographics** | | | | |
| Age (years) | **0.83 (0.11), < 0.01** | **0.87 (0.11), < 0.01** | **0.85 (0.12), < 0.01** | **0.47 (0.11), < 0.01** |
| Male (vs F) | 1.00 (0.73), 0.17 | 1.05 (0.73), 0.15 | 1.07 (0.73), 0.15 | 0.52 (0.65), 0.43 |
| Black (vs W) | 0.38 (0.72), 0.59 | 0.47 (0.72), 0.51 | 0.63 (0.73), 0.39 | 0.97 (0.65), 0.14 |
| **Within 1st hour** | | | | |
| HR avg | **0.05 (0.02), < 0.01** | **0.06 (0.02), < 0.01** | **0.09 (0.04), 0.05*** | **0.08 (0.04), 0.05*** |
| RR avg | 0.00 (0.04), 0.99 | 0.02 (0.04), 0.68 | -0.06 (0.09), 0.50 | -0.06 (0.08), 0.51 |
| SpO2 avg | **-0.39 (0.14), < 0.01** | **-0.37 (0.14), < 0.01** | -0.24 (0.21), 0.25 | -0.12 (0.19), 0.53 |
| Magnesium | | 0.16 (3.34), 0.96 | 0.35 (3.65), 0.92 | 0.46 (3.22), 0.89 |
| Epinephrine | | -1.95 (2.81), 0.49 | -1.54 (3.91), 0.69 | -1.04 (3.46), 0.76 |
| Alb 5mg | | -1.35 (8.14), 0.87 | -0.26 (8.89), 0.98 | -0.01 (7.87), 1.00 |
| Alb 10mg | | -2.64 (1.38), 0.06 | 0.77 (2.15), 0.72 | 0.89 (1.89), 0.64 |
| Alb 15mg | | -1.27 (0.76), 0.09 | -0.02 (1.08), 0.98 | 0.23 (0.96), 0.81 |
| **Within 2 hours** | | | | |
| HR avg | | | -0.04 (0.05), 0.37 | -0.08 (0.07), 0.21 |
| RR avg | | | 0.10 (0.11), 0.34 | -0.04 (0.13), 0.77 |
| SpO2 avg | | | -0.18 (0.22), 0.40 | -0.01 (0.24), 0.96 |
| Magnesium | | | -0.23 (1.66), 0.89 | -1.16 (1.69), 0.49 |
| Epinephrine | | | -0.46 (2.82), 0.87 | -2.10 (3.31), 0.53 |
| Alb 5mg | | | -0.75 (3.67), 0.84 | 0.38 (5.70), 0.95 |
| Alb 10mg | | | **-4.19 (1.83), 0.02** | -4.02 (2.65), 0.13 |
| Alb 15mg | | | -1.99 (1.11), 0.07 | -0.74 (1.36), 0.59 |
| **Within 4 hours** | | | | |
| HR avg | | | | 0.02 (0.06), 0.72 |
| RR avg | | | | 0.14 (0.11), 0.20 |
| SpO2 avg | | | | -0.37 (0.24), 0.12 |
| Magnesium | | | | 1.68 (1.05), 0.11 |
| Epinephrine | | | | 1.57 (2.28), 0.49 |
| Alb 5mg | | | | 0.02 (4.71), 1.00 |
| Alb 10mg | | | | -0.19 (2.21), 0.93 |
| Alb 15mg | | | | -1.00 (1.26), 0.42 |

Table shows the effect estimates (β regression coefficients) and standard errors (SE) for each variable. Model A indicates the model described in Supplemental Table 1 (total LOS for children admitted without PICU) and Supplemental Figure 2C. Alb: Albuterol. *P < 0.05.

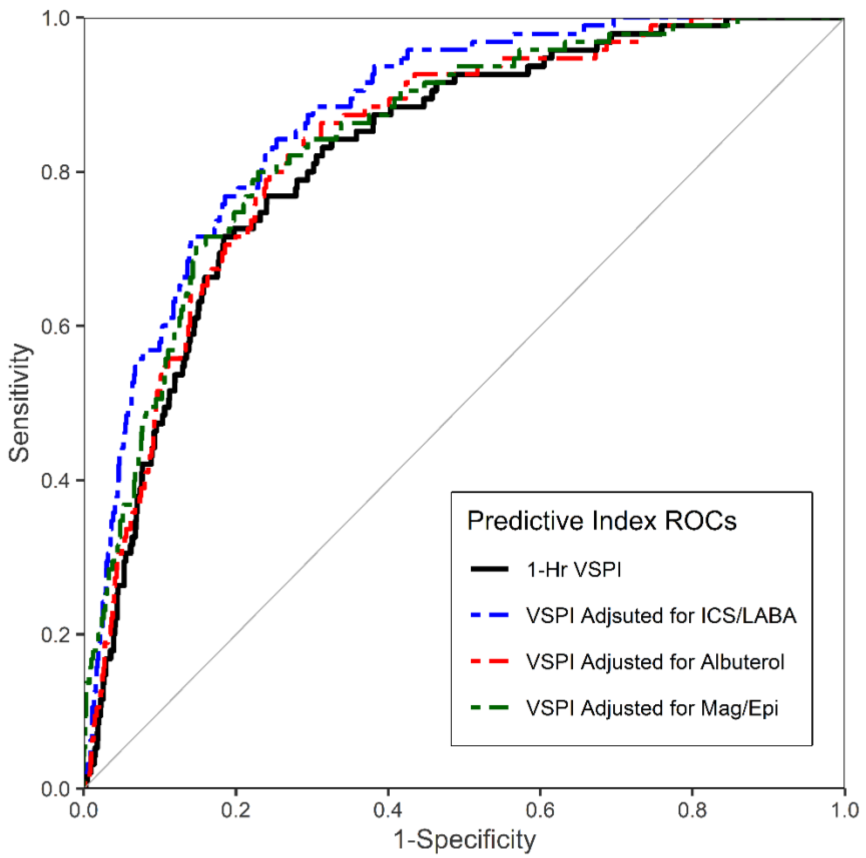


Supplemental Figure 1. Receiver-Operator Curves (ROC) of the predictive index using demographics and 1st hour vital signs to predict PICU admission (black), compared to the ROCs of the same vital signs predictive index (VSPI) adjusted for ICS/LABA use while inpatient (blue), albuterol administration in the ED (red), magnesium or epinephrine administration in the ED (green).

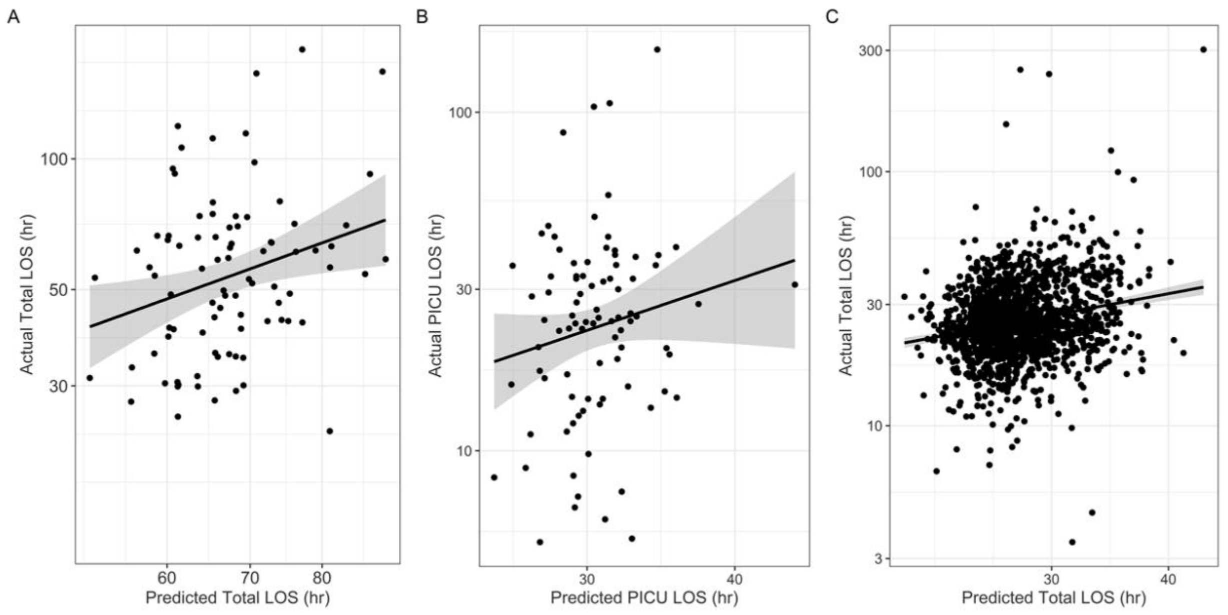


Supplemental Figure 2. Predicted vs Actual LOS. (A, left) Total LOS (β=0.27, p=0.01, $R^2$=0.07) and (B, middle) PICU LOS (β=0.04, p=0.23, $R^2$=0.02) among those admitted to the PICU with an acute asthma exacerbation. (C, right) Predicted vs Actual Total LOS among those admitted to the hospital without PICU (β=0.22, p<0.01, $R^2$=0.05). Scales are logarithmic. Trendlines indicate best fit of linear regression model with grey margin indicating standard error.
